# Excess mortality in Cyprus during the COVID-19 pandemic and its lack of association with vaccination rates

**DOI:** 10.1101/2022.08.05.22278487

**Authors:** Theodore Lytras, Maria Athanasiadou, Anna Demetriou, Despina Stylianou, Alexandros Heraclides, Olga Kalakouta

## Abstract

**Background:** It has been claimed that COVID-19 vaccination is associated with excess mortality during the COVID-19 pandemic, a claim that contributes to vaccine hesitancy. We examined whether all-cause mortality has actually increased in Cyprus during the first two pandemic years, and whether any increases are associated with vaccination rates.

**Methods:** We calculated weekly excess mortality for Cyprus between January 2020 and June 2022, overall and by age group, using both a Distributed Lag Nonlinear Model (DLNM) adjusted for mean daily temperature, and the EuroMOMO algorithm. Excess deaths were regressed on the weekly number of confirmed COVID-19 deaths and on weekly first-dose vaccinations, also using a DLNM to explore the lag-response dimension.

**Results:** 552 excess deaths were observed in Cyprus during the study period (95%CI: 508–597) as opposed to 1306 confirmed COVID-19 deaths. No association between excess deaths and vaccination rates was found overall and for any age group except 18-49 years, among whom 1.09 excess deaths (95%CI: 0.27–1.91) per 10,000 vaccinations were estimated during the first 8 weeks post-vaccination. However, detailed cause-of-death examination identified just two such deaths potentially linked to vaccination, therefore this association is spurious and attributable to random error.

**Conclusions:** Excess mortality was moderately increased in Cyprus during the COVID-19 pandemic, primarily as a result of laboratory-confirmed COVID-19 deaths. No relationship was found between vaccination rates and all-cause mortality, demonstrating the excellent safety profile of COVID-19 vaccines.

## Introduction

During the COVID-19 pandemic, scientists and the public have been using reported COVID-19 daily deaths to monitor the course of the pandemic [1]. These counts are commonly based on laboratory confirmation of SARS-CoV-2 infection, thereby excluding deaths due to COVID-19 that for any reason are not tested for SARS-CoV-2 (especially some out-of-hospital deaths) [2]. As a result, they represent an incomplete account of COVID-19 mortality, which in turn is more reliably captured by “excess mortality”, the increase in all-cause deaths over those expected based on historic trends [3]. On the other hand, excess mortality presents challenges of its own such as the difficulty to distinguish direct and indirect effects of the pandemic, as well as its sensitivity to the modelling choices used to estimate expected deaths [4,5].

The complexity in analyzing and interpreting all-cause mortality data has laid the door open to abuse and misrepresentation by anti-vaccine activists and conspiracy theorists, trying to link COVID-19 vaccination to increased mortality [6] despite overwhelming direct [7] and indirect [8] evidence on the effectiveness of vaccination in preventing COVID-19 deaths. In Cyprus, one such claim concerns an increase in all-cause mortality “concurrent” with the COVID-19 vaccination campaign; this claim has been widely reproduced in the media [9,10] and even emulated by others abroad [11], thereby contributing to vaccine hesitancy. Recently a published paper has alleged increased all-cause mortality in Cyprus and temporally linked it to mass COVID-19 vaccination [12], a claim again reproduced in the media to stoke vaccine skepticism [13].

In this paper we set out to examine (a) whether all-cause mortality in the Republic of Cyprus has actually increased during the COVID-19 pandemic, and (b) whether any excess mortality is temporally associated with COVID-19 vaccination, using rigorous statistical methodology and analyzing both the entire population and specific age groups. Appropriately exploring the temporal dimension is critical to ascribing causality: if vaccination does increase the risk of death, it should do so by a consistent amount at particular time offsets, whereas a transient and isolated mortality excess has to be attributed to other causes.

## Methods

### Data sources, study period and definitions

For all-cause and (laboratory-confirmed) COVID-19 deaths in Cyprus by date and age we used anonymized official data collected by the Health Monitoring Unit of the Cyprus Ministry of Health until ISO week 26/2022. For COVID-19 vaccinations in Cyprus by week and age group we used the data reported by the European Centre for Disease Prevention and Control [14], and from the National Oceanic and Atmospheric Administration (NOAA) [15] we downloaded mean daily temperature data and calculated a population-weighted daily mean across all weather stations in Cyprus. The study period was from week 01/2020 to 22/2022 (i.e. four weeks prior to data retrieval) in order to minimise the effect of delayed reporting of deaths. To calculate baseline expected mortality we used mortality and temperature data up to 10 years prior (from week 01/2010); according to these historical data, only 6.4% of deaths are registered later than four weeks from the date of death. We ran four sets of analyses, one for the entire population of the Republic of Cyprus, and for three age groups: 18-49, 50-69 and ≥70 years.

### Estimating excess mortality

For each set, we ran a two-stage analysis. At the first stage we estimated daily and weekly excess mortality adjusted for trend, seasonality and mean daily temperature, using a Distributed Lag Nonlinear Model (DLNM) [16] of quasi-Poisson family with log link, similar to a model previously employed for estimating influenza-attributable mortality in Greece [17]. Our model included a linear trend, a periodic B-spline for seasonality with three equidistant knots for day of the year, and a cross-basis for temperature whose exposure-response curve was modelled with a quadratic B-spline with three internal knots at the 10th, 75th and 90th percentile of temperature values, and its lag-response with a natural cubic spline with three knots equidistant in the log scale [17]. The model was fitted to the mortality and temperature data between weeks 01/2010 and 52/2019, then used to calculate predicted deaths (and thus excess deaths, as observed minus predicted) for the study period (weeks 01/2020 to 22/2022). As a sensitivity analysis, we additionally employed the well-known EuroMOMO algorithm to calculate excess deaths for the same study period, based on the mortality of the preceding five years; importantly, EuroMOMO does not take account of temperature data in the calculation, but only a linear trend and a sinus term for seasonality [18].

### Analyzing the association between excess mortality and vaccination

At the second-stage analysis, we ran a linear regression of the weekly number of excess deaths for the study period, on the weekly number of laboratory-confirmed COVID-19 deaths and on a cross-basis (variable transformation [16]) for the weekly new first-dose COVID-19 vaccinations, thereby modelling the association between vaccinations and excess deaths at multiple time lags. We used two alternative cross-basis matrices, one “short” and one “long”, to examine potential short-term and long-term effects of vaccination on mortality; in both cases the exposure-response relationship was set to be linear without an intercept, whereas the lag-response relationship for the “short” version was set as stratified with one indicator per week up to 8 weeks, and for the “long” version as following a natural cubic spline with three equidistant knots up to 52 weeks. The “short” cross-basis matrix assumes vaccination may influence the risk of death only for the first two months after vaccination (thereby including the month after the second vaccine dose, for mRNA vaccines), whereas the “long” version extends this to an entire year after vaccination.

To guard against overfitting, both models were compared to a “null model” (containing only weekly COVID-19 deaths as exposure variable) using a Likelihood Ratio Test and the Akaike Information Criterion; provided the inclusion of vaccination resulted in a significantly improved model fit (using both criteria), the resulting association between excess mortality and vaccination was examined quantitatively (overall, i.e. across all lags) and qualitatively (i.e. its shape along the lag-response dimension).

In case a “short”-version model showed both an improved fit and a statistically significant overall association between excess mortality and COVID-19 vaccination, we investigated further by (a) determining how many deaths actually occurred during 2021 (most recent year for which detailed individual data were available) during the first 8 weeks from a person’s first-dose vaccination, (b) descriptively analyzing the causes of death for these individuals, and (c) comparing with the model-predicted number of excess deaths during 2021, given the total vaccine doses administered in the first 44 weeks of 2021. If the estimated association were true, we would expect to observe a number of deaths with causes potentially linked to COVID-19 vaccination, approximately in the range predicted by the model; if not, the association has to be non-causal and attributed to random error.

All statistical analyses were performed with the R statistical environment, version 4.2.1 [19]. Data are available as supplementary material with this article.

## Results

The estimated excess all-cause mortality, observed COVID-19 deaths and number of first-dose vaccinations, overall and by age group, are illustrated on Table 1 and Figure 1. As expected, excess mortality estimates with EuroMOMO are mostly higher than with the DLNM, since the former does not account for the anticipated effect of high or low temperatures on mortality. Vaccination coverage over the study period was higher with increasing age, as vaccination roll-out was staggered by age group. Most weeks with excess deaths also had many COVID-19 deaths (Figure 1) and overall excess mortality did not exceed overall COVID-19 deaths with the exception of the 18-49 age group (Table 1).

**Table 1:**
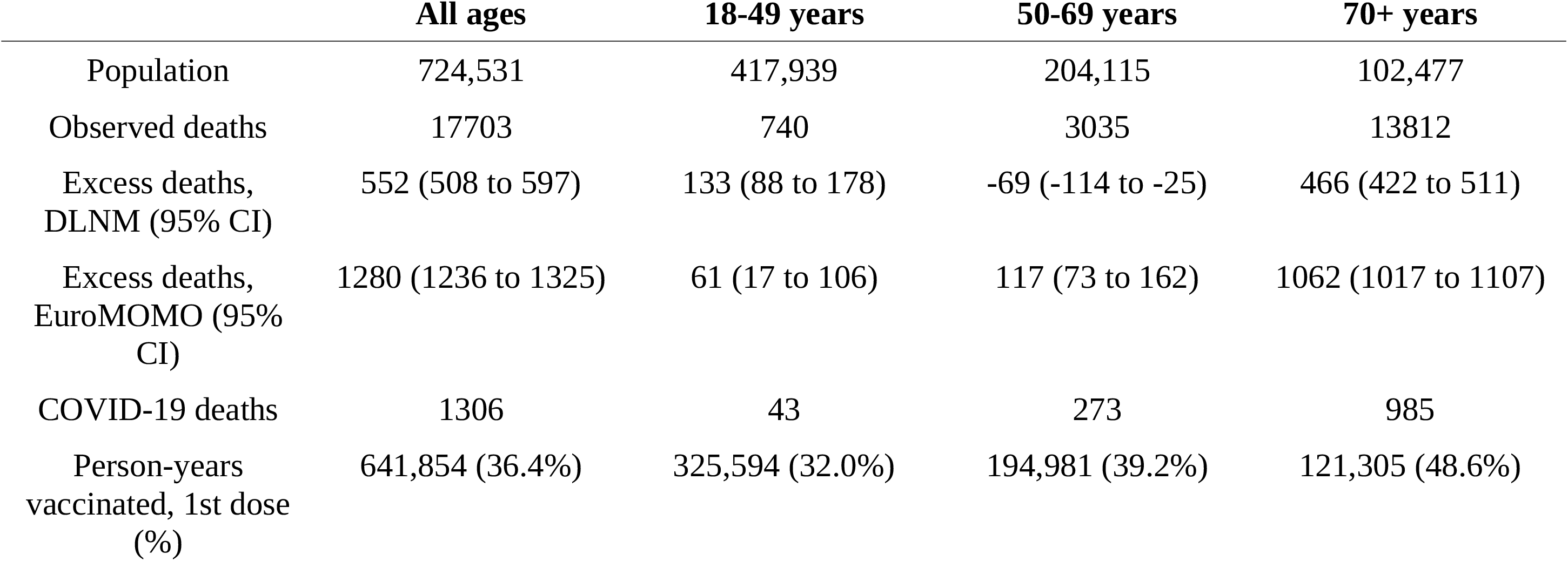
Observed and excess mortality, COVID-19 deaths and vaccination coverage in Cyprus, overall and by age group, over the study period (week 01/2020 to 22/2022)

|  | **All ages** | **18-49 years** | **50-69 years** | **70+ years** |
| --- | --- | --- | --- | --- |
| Population | 724,531 | 417,939 | 204,115 | 102,477 |
| Observed deaths | 17703 | 740 | 3035 | 13812 |
| Excess deaths, DLNM (95% CI) | 552 (508 to 597) | 133 (88 to 178) | -69 (-114 to -25) | 466 (422 to 511) |
| Excess deaths, EuroMOMO (95% CI) | 1280 (1236 to 1325) | 61 (17 to 106) | 117 (73 to 162) | 1062 (1017 to 1107) |
| COVID-19 deaths | 1306 | 43 | 273 | 985 |
| Person-years vaccinated, 1st dose (%) | 641,854 (36.4%) | 325,594 (32.0%) | 194,981 (39.2%) | 121,305 (48.6%) |

**Figure 1:**
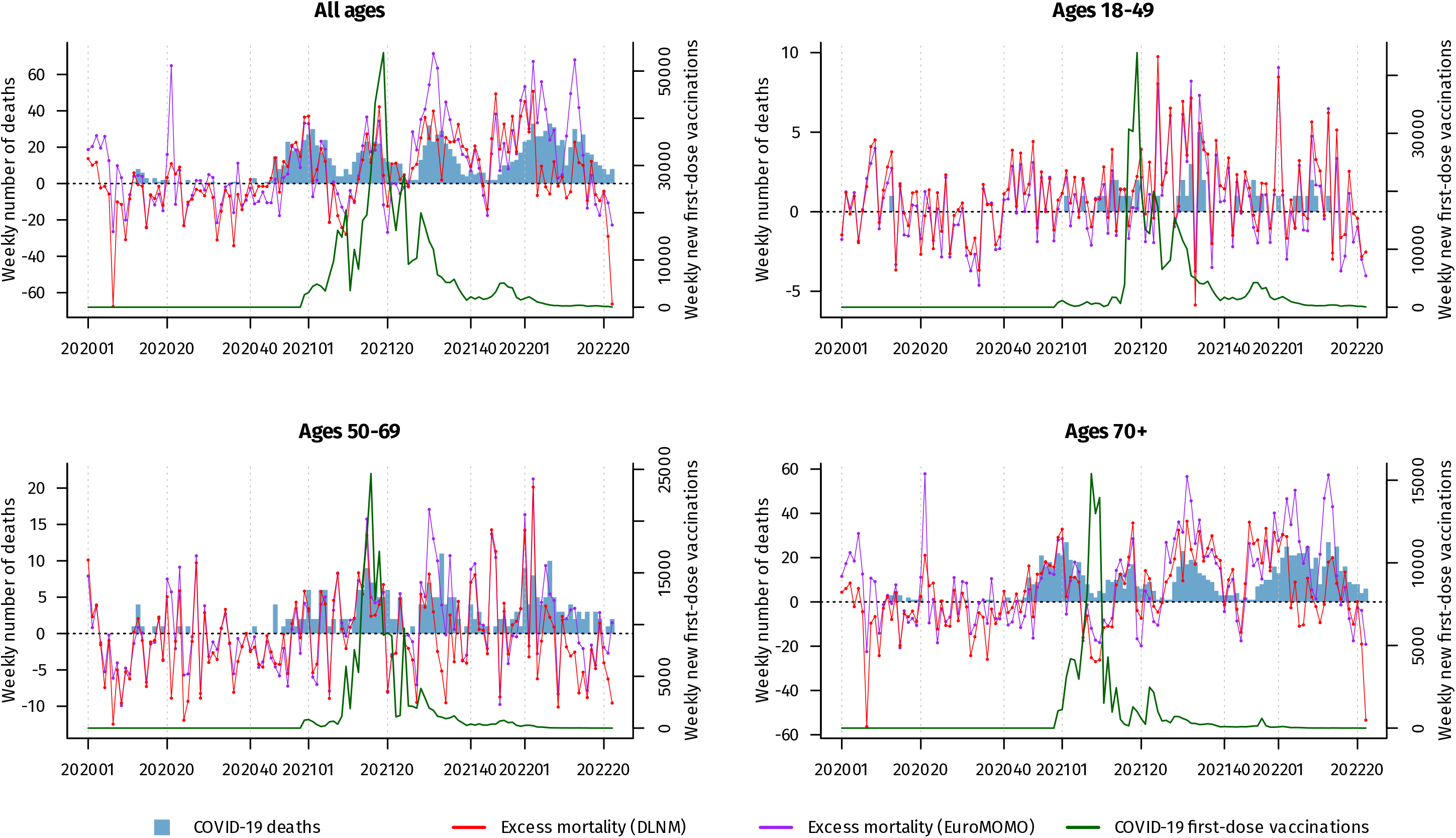
Weekly excess mortality, COVID-19 deaths and first-dose vaccinations in Cyprus, overall and by age group, over the study period (week 01/2020 to 22/2022)

The comparison of second-stage model fits is shown on Table 2; statistically significant improved fits (in bold) indicate a potential association between vaccination and excess mortality. These associations are illustrated in Figure 2 (full results for all models in Supplementary Figure 1) both overall (across all lags) and across the lag-response dimension. For all ages, 50-69 and 70+ years the overall associations did not reach statistical significance; in addition, the lag-response curves were erratic (indicating both mortality excesses and deficits at different lag times) in a manner that is unlikely to indicate a causal association. On the other hand, for the 18-49 years age group a small short-term association was estimated, with 1.09 (95% CI: 0.27 to 1.91) excess deaths per 10,000 vaccinations during the first 8 weeks post-vaccination. This estimate remained in the sensitivity analysis using EuroMOMO (0.84 deaths, 95% CI: 0.04 to 1.64, Supplementary Figure 1), whereas when using a long (52 weeks) cross-basis for vaccination the association became non-significant, though still positive for the first few weeks.

**Table 2:** Second-stage model fits, short vs long cross-basis for vaccination, overall and by age group.

|  | Null model | Short cross-basis | Long cross-basis |
| --- | --- | --- | --- |
| *All ages* |  |  |  |
| AIC (DLNM) | 1090.7 | 1096.3 | **1059.7** |
| LRT p-value (DLNM) | – | 0.27 | **<0.001** |
| AIC (EuroMOMO) | 1104.3 | **1097.6** | **1074.5** |
| LRT p-value (EuroMOMO) | – | **0.004** | **<0.001** |
| *Ages 18-49* |  |  |  |
| AIC (DLNM) | 581.9 | **577.5** | 584.9 |
| LRT p-value (DLNM) | – | **0.009** | 0.24 |
| AIC (EuroMOMO) | 578.9 | **570.9** | 581.1 |
| LRT p-value (EuroMOMO) | – | **0.002** | 0.18 |
| *Ages 50-69* |  |  |  |
| AIC (DLNM) | 797.3 | 808.3 | **791.8** |
| LRT p-value (DLNM) | – | 0.79 | **0.008** |
| AIC (EuroMOMO) | 779.7 | 789.2 | 782.6 |
| LRT p-value (EuroMOMO) | – | 0.63 | 0.22 |
| *Ages 70+* |  |  |  |
| AIC (DLNM) | 1054.5 | 1054.6 | **1011.7** |
| LRT p-value (DLNM) | – | 0.049 | **<0.001** |
| AIC (EuroMOMO) | 1064.9 | 1067.6 | **1049.4** |
| LRT p-value (EuroMOMO) | – | 0.11 | **<0.001** |

AIC = Akaike Information Criterion (lower indicates better fit), LRT = Likelihood Ratio Test. Bold face indicates a statistically significant and better model fit.

**Figure 2:**
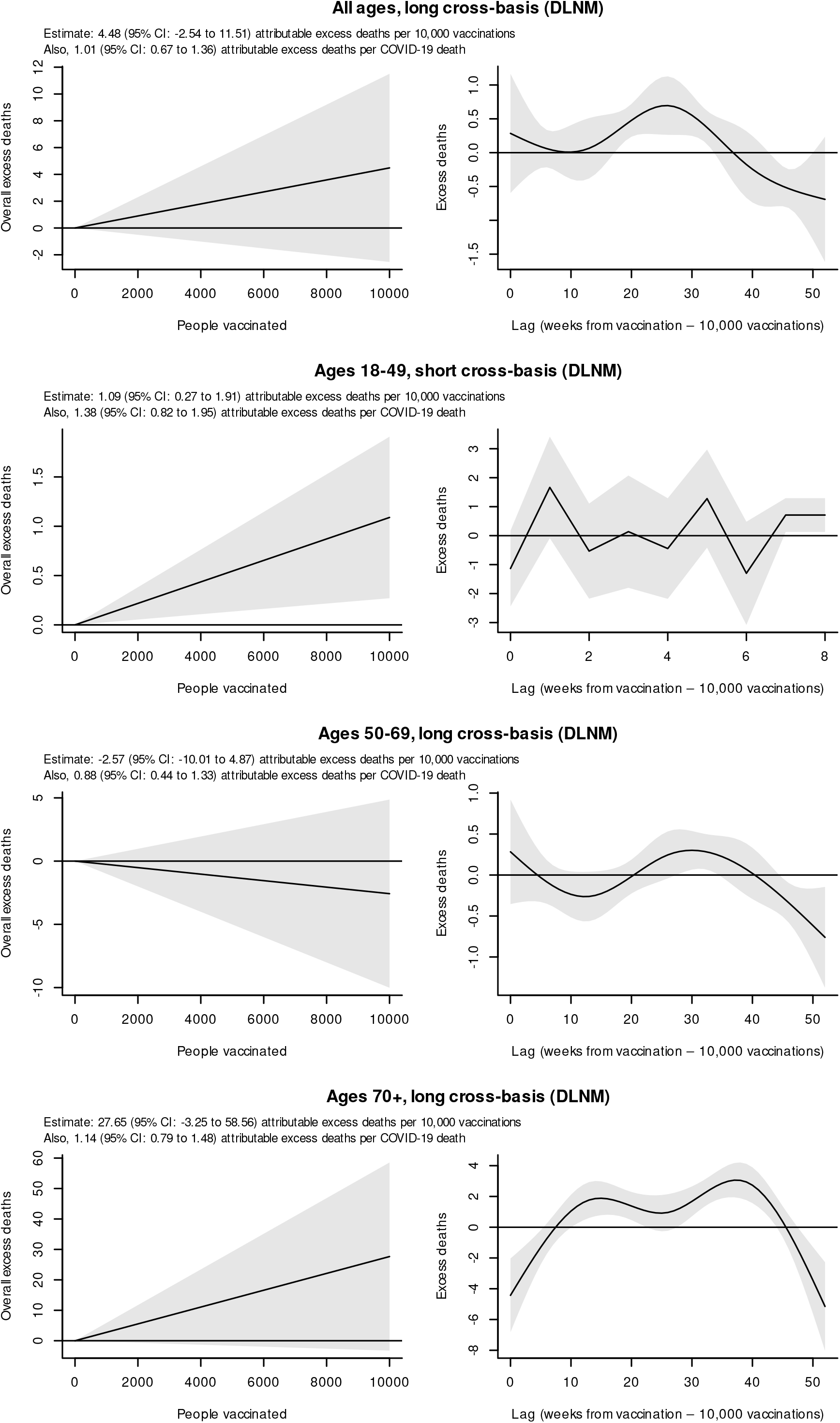
Associations between excess mortality, COVID-19 deaths and first-dose vaccinations in Cyprus (age groups with statistically significant and improved model fits)

Under the hypothetical assumption of causality for this observed association and given that 313,032 people aged 18-49 were vaccinated until week 44/2021, we would expect during 2021 a total of 34 deaths (95% CI: 8 to 60) in this age group to be attributable to COVID-19 vaccination up to 8 weeks before death. This occurs in a context of 34 confirmed COVID-19 deaths and 337 all-cause deaths in this age group during 2021. We investigated the association further, by going back to the Republic of Cyprus Ministry of Health data and querying how many persons aged 18-49 during the study period *actually* passed away up to 8 weeks after receiving their first vaccine dose. We found 18 such persons with a median age of 41 years, of whom: 5 died of trauma, 1 due to severe COVID-19, 5 due to pre-existing malignancy, 1 due to chronic liver cirrhosis, 1 due to a stroke and 3 due to myocardial infarction (2 of whom had previously diagnosed coronary artery disease). Additionally there was one death with diagnosed Vaccine-Induced Thrombotic Thrombocytopenia (VITT) [20] and another due to a subarachnoid haemorrage [21], both in women and both following receipt of the ChAdOx1 nCov-19 vaccine. Given that only these two deaths were vaccination-related, rather than the 34 expected based on our analysis, we conclude the association observed between vaccination and excess mortality in the 18-49 year age group is false, attributable to type I random error, and therefore non-causal. For all other age groups, no association between COVID-19 vaccination and all-cause mortality was observed (Figure 2).

In all models and for all age groups, the ratio between confirmed COVID-19 and excess deaths was always around one (95% CIs including one) thereby indicating no substantial under-or over-ascertainment of COVID-19 deaths, given that laboratory-confirmed deaths are likely the best proxy indicator for the impact of coronavirus on population mortality, and assuming relatively stable ascertainment over time.

## Discussion

Our study indicates that all-cause mortality in Cyprus was only moderately increased from 2020 to 2022, and this increase is largely accounted for by laboratory-confirmed COVID-19 deaths. In all but one analysis, no evidence of an association between COVID-19 vaccination and excess mortality was found, thereby disproving rumors and theories about large increases in all-cause mortality as a result of vaccination. Indeed vaccines are by far the most effective way to protect against COVID-19 severe disease and death, and have saved countless lives during the current pandemic [7].

In the 18-49 years age group a weak but statistically significant association with excess mortality was observed only for the first 8 weeks after COVID-19 vaccination. However, after reviewing the deaths that actually occured in this timeframe and age group, we found many fewer than would be expected if the association were true; therefore causality was disproven and the association attributed to type I random error. This is in line with past studies and current knowledge about COVID-19 vaccines; although the short-term risks of myocarditis and pericarditis following mRNA COVID-19 vaccination are well-described [22], they are much rarer (only a few cases per 100,000 vaccinations) and largely mild clinically. Furthermore, a report from England found no increase in excess mortality among young adults in relation to COVID-19 vaccination [23]. Our analysis provides additional confirmation about the safety of COVID-19, including in younger adults.

Although not an objective of our study, we identified one diagnosed and one suspect VITT death after vaccination with ChAdOx1 nCov-19 in persons aged 18-49 years, out of 39,657 doses administered in this age group. This rate is within the range reported in the literature, though still on the high side [24].

It has been argued that COVID-19 deaths can be not just underestimated due to lack of testing, but also potentially overestimated in case of excessive testing if people are found positive to SARS-CoV-2 incidentally around the time of their death [25]. Our analysis found approximately one-to-one ratio between weekly confirmed COVID-19 deaths and excess deaths, both overall and in each age group; this shows that surveillance in Cyprus reliably detected most if not all COVID-19 deaths, and alleviates concerns about potential overascertainment. Furthermore, and considering this one-to-one ratio, if we subtract confirmed COVID-19 deaths from all-cause deaths then we have a mortality deficit (rather than excess) both overall and for the 50-69 and 70+ age groups. This shows COVID-19 had little to no indirect impact on population mortality in Cyprus, in line with previous studies [26], and suggests measures employed in the country to control the pandemic were overall fairly successful in safeguarding public health.

Finally, our results illustrate the importance of having a solid framework to estimate expected mortality for a time period, in order to obtain credible excess mortality estimates (since excess deaths are observed minus expected). Past years’ annual death counts are usually insufficient for analysis, as they ignore both trend, seasonality and historical excesses due to other causes. In addition, when the goal is to infer about specific factors affecting excess mortality (as was the case with our study) it is beneficial to adjust for unrelated major determinants of mortality such as ambient temperature when estimating expected death counts. Adjusting for ambient temperature with our DLNM model in most cases produced substantially lower excess mortality estimates compared to the EuroMOMO algorithm (which does not make such an adjustment), even though the second-stage results we obtained were overall similar with both approaches.

In conclusion, our study showed only moderately increased all-cause mortality in Cyprus during the COVID-19 pandemic; this excess is largely accounted for by confirmed COVID-19 deaths, and importantly has no association with vaccination rates either overall or by age group, both in a short (8 weeks post-vaccination) and in a long (1 year post-vaccination) timeframe. Our results reconfirm the excellent safety of COVID-19 vaccination and will hopefully help in promoting vaccine uptake. Finally, the analytical approach presented here can be useful for other countries in the context of vaccine safety monitoring, in order to detect potential excess mortality signals and drive further investigation if needed.

## Supporting information

Study data

## Data Availability

The data analyzed by this study are contained in the manuscript as supplementary material

## Key Points

- It has been claimed that mass COVID-19 vaccination is associated with increased all-cause mortality, a claim that contributes to vaccine hesitancy
- We calculated excess mortality in Cyprus during the pandemic period, and examined its association with COVID-19 vaccination rates across multiple time lags (up to a year)
- Excess mortality was largely accounted for by laboratory-confirmed COVID-19 deaths
- No short- or long-term association between excess mortality and COVID-19 vaccination was found
- Findings reconfirm the safety of COVID-19 vaccines and assist in combating anti-vaccine claims and vaccine hesitancy, thereby promoting vaccine uptake

## Figure Legends

**Supplementary Figure 1:** Associations between excess mortality, COVID-19 deaths and first-dose vaccinations in Cyprus (all age groups, all models)

