## Supplementary Figure 1 for "Excess mortality in Cyprus during the COVID-19 pandemic and its lack of association with vaccination rates"

### All ages, short cross-basis (DLNM)

Estimate: 4.19 (95% CI: 0.43 to 7.96) attributable excess deaths per 10,000 vaccinations
Also, 0.87 (95% CI: 0.57 to 1.18) attributable excess deaths per COVID-19 death

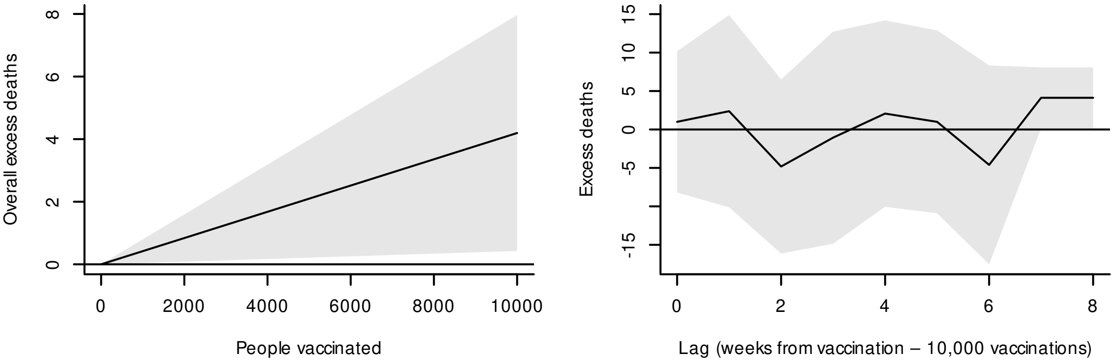


### All ages, short cross-basis (EuroMOMO)

Estimate: 1.36 (95% CI: -2.43 to 5.15) attributable excess deaths per 10,000 vaccinations
Also, 1.33 (95% CI: 1.02 to 1.64) attributable excess deaths per COVID-19 death

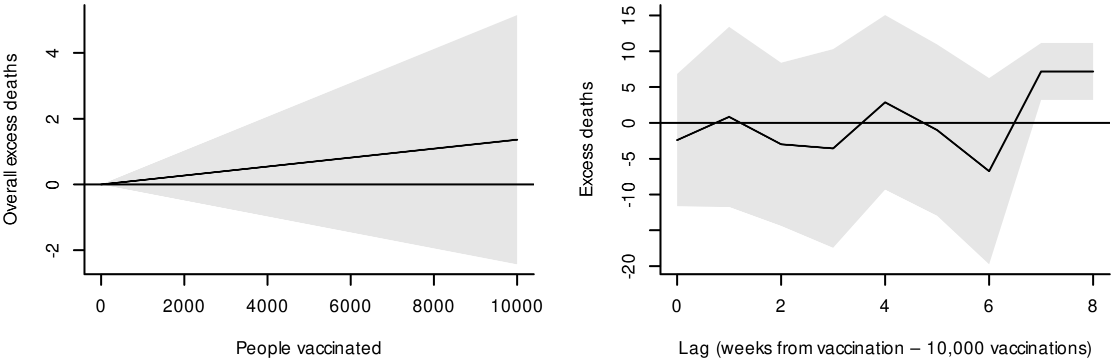


### All ages, long cross-basis (DLNM)

Estimate: 4.48 (95% CI: -2.54 to 11.51) attributable excess deaths per 10,000 vaccinations
Also, 1.01 (95% CI: 0.67 to 1.36) attributable excess deaths per COVID-19 death

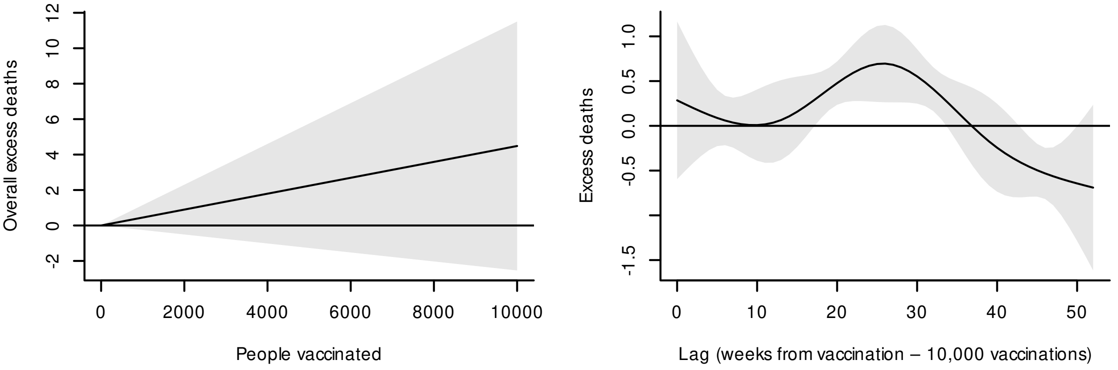


### All ages, long cross-basis (EuroMOMO)

Estimate: 6.55 (95% CI: -0.89 to 14.00) attributable excess deaths per 10,000 vaccinations
Also, 0.93 (95% CI: 0.56 to 1.29) attributable excess deaths per COVID-19 death

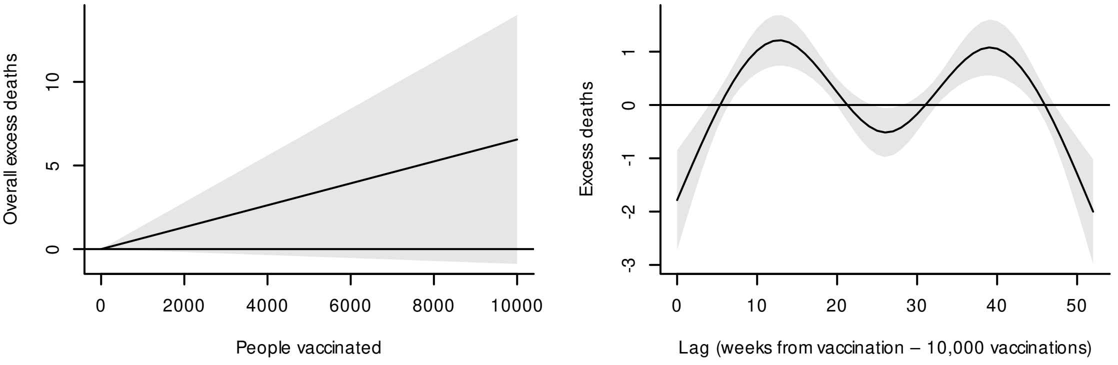

### Ages 18-49 ages, short cross-basis (DLNM)

Estimate: 1.09 (95% CI: 0.27 to 1.91) attributable excess deaths per 10,000 vaccinations
Also, 1.38 (95% CI: 0.82 to 1.95) attributable excess deaths per COVID-19 death

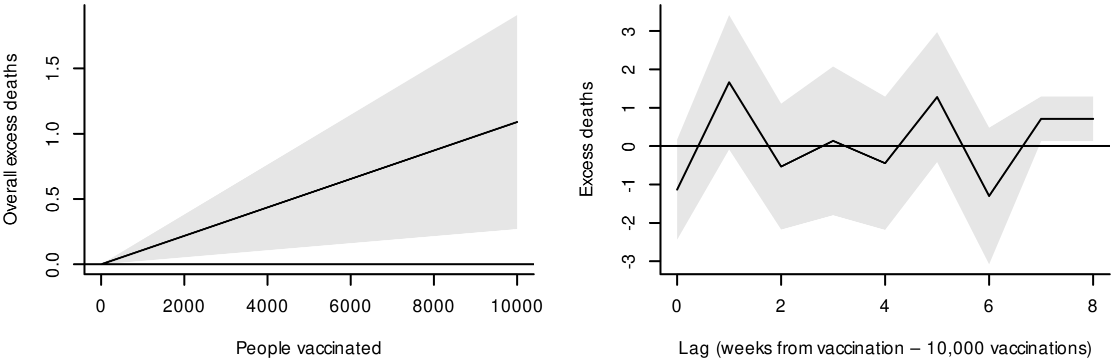


### Ages 18-49 ages, short cross-basis (EuroMOMO)

Estimate: 0.86 (95% CI: 0.06 to 1.65) attributable excess deaths per 10,000 vaccinations
Also, 1.66 (95% CI: 1.11 to 2.21) attributable excess deaths per COVID-19 death

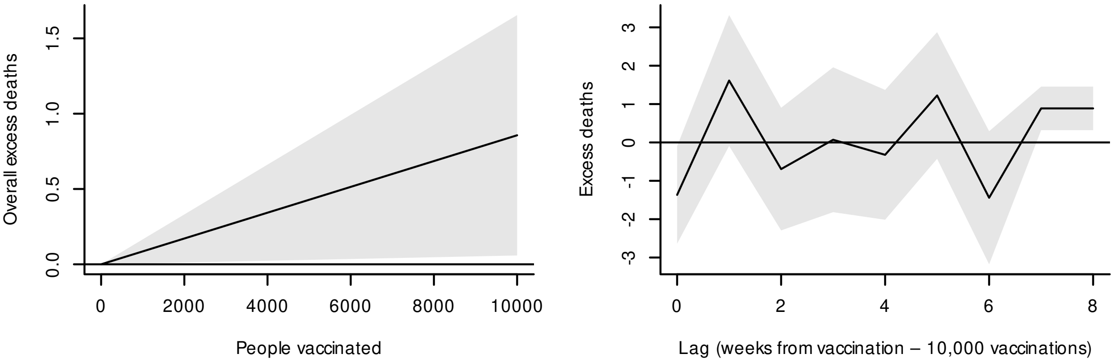


### Ages 18-49 ages, long cross-basis (DLNM)

Estimate: 1.25 (95% CI: -0.36 to 2.85) attributable excess deaths per 10,000 vaccinations
Also, 1.12 (95% CI: 0.51 to 1.73) attributable excess deaths per COVID-19 death

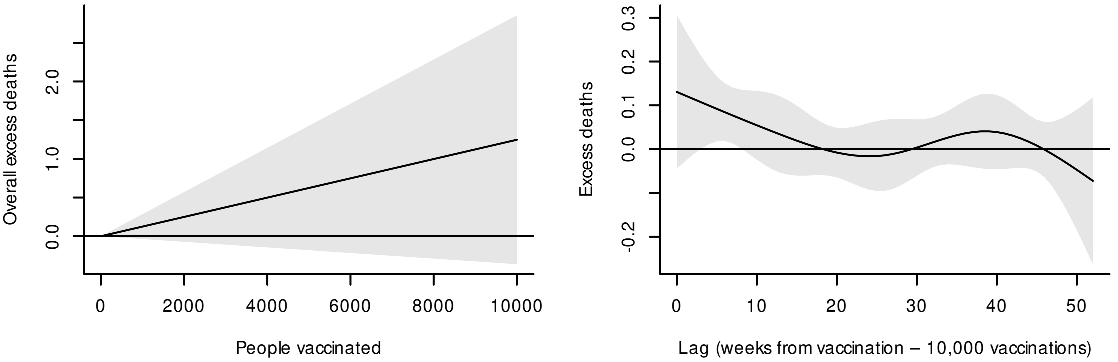


### Ages 18-49 ages, long cross-basis (EuroMOMO)

Estimate: 0.61 (95% CI: -0.98 to 2.19) attributable excess deaths per 10,000 vaccinations
Also, 1.28 (95% CI: 0.68 to 1.88) attributable excess deaths per COVID-19 death

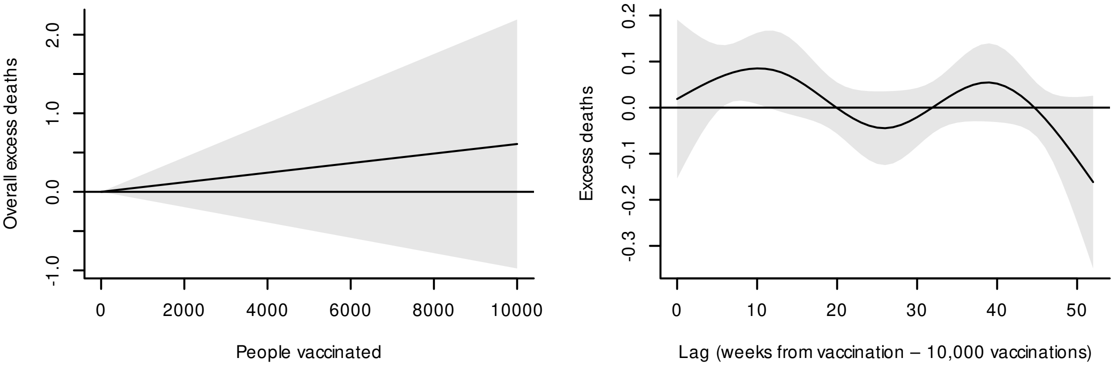

**Ages 50-69 ages, short cross-basis (DLNM)**

Estimate: 0.84 (95% CI: -2.68 to 4.36) attributable excess deaths per 10,000 vaccinations
Also, 0.75 (95% CI: 0.31 to 1.19) attributable excess deaths per COVID-19 death

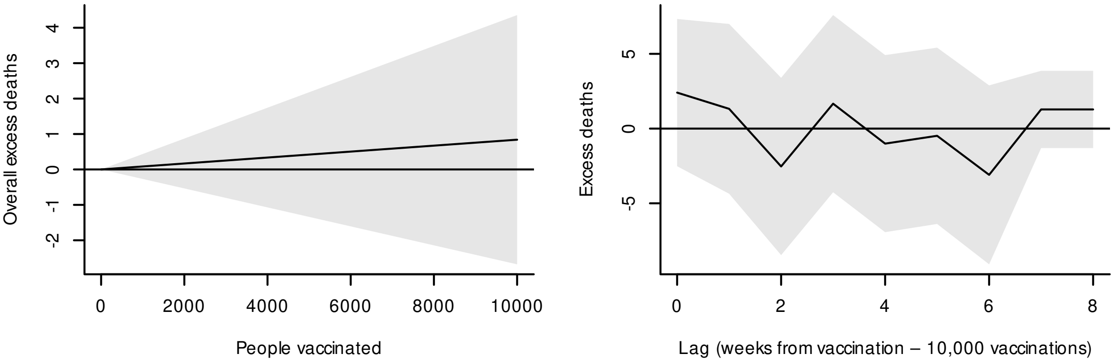


**Ages 50-69 ages, short cross-basis (EuroMOMO)**

Estimate: 0.03 (95% CI: -3.23 to 3.29) attributable excess deaths per 10,000 vaccinations
Also, 1.21 (95% CI: 0.80 to 1.61) attributable excess deaths per COVID-19 death

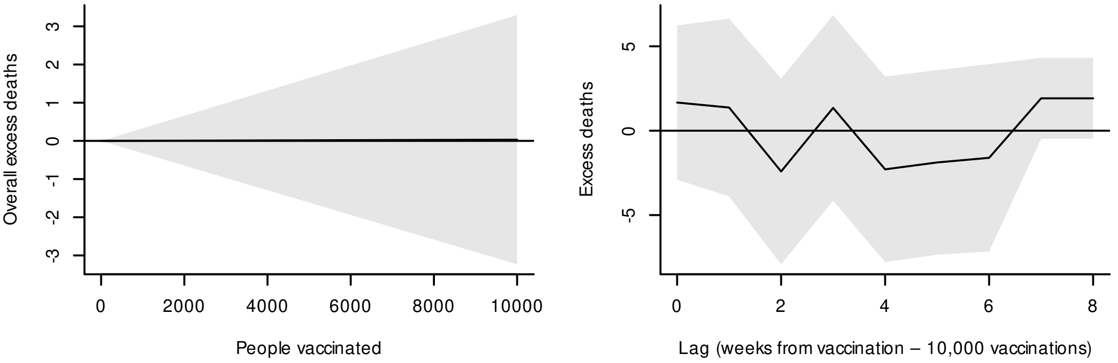


**Ages 50-69 ages, long cross-basis (DLNM)**

Estimate: -2.57 (95% CI: -10.01 to 4.87) attributable excess deaths per 10,000 vaccinations
Also, 0.88 (95% CI: 0.44 to 1.33) attributable excess deaths per COVID-19 death

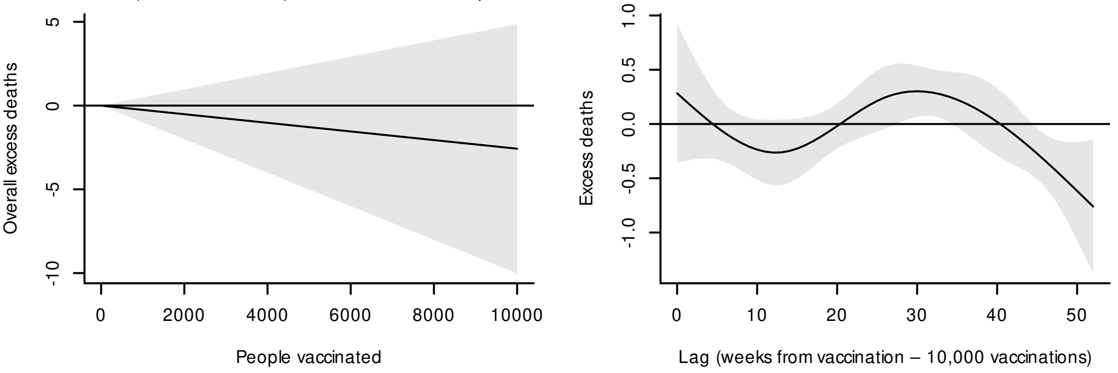


**Ages 50-69 ages, long cross-basis (EuroMOMO)**

Estimate: 0.88 (95% CI: -6.30 to 8.05) attributable excess deaths per 10,000 vaccinations
Also, 1.15 (95% CI: 0.72 to 1.59) attributable excess deaths per COVID-19 death

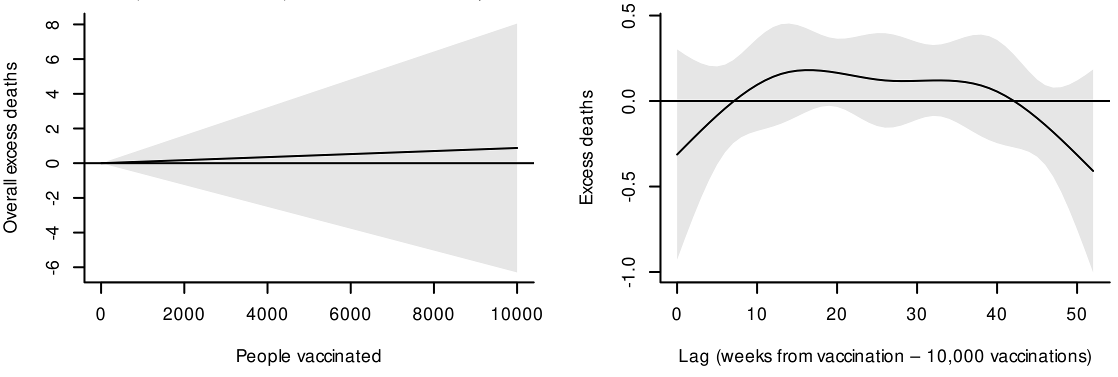

**Ages 70+ ages, short cross-basis (DLNM)**

Estimate: -14.66 (95% CI: -29.68 to 0.36) attributable excess deaths per 10,000 vaccinations
Also, 0.90 (95% CI: 0.58 to 1.23) attributable excess deaths per COVID-19 death

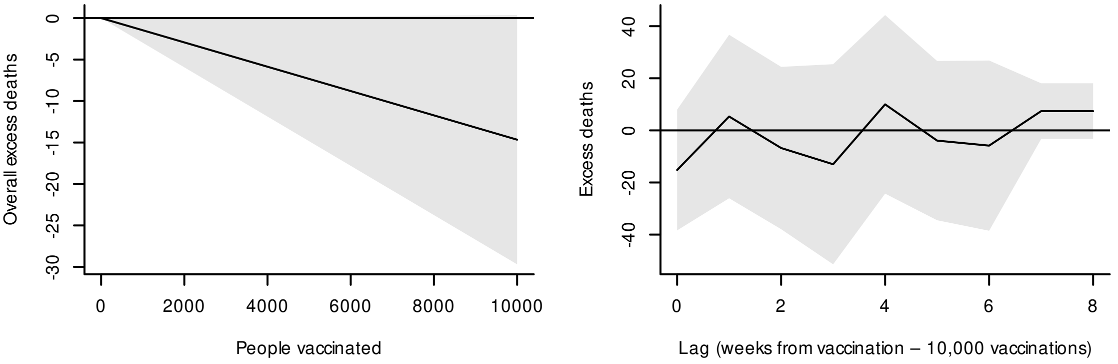


**Ages 70+ ages, short cross-basis (EuroMOMO)**

Estimate: -19.40 (95% CI: -35.21 to -3.59) attributable excess deaths per 10,000 vaccinations
Also, 1.28 (95% CI: 0.94 to 1.63) attributable excess deaths per COVID-19 death

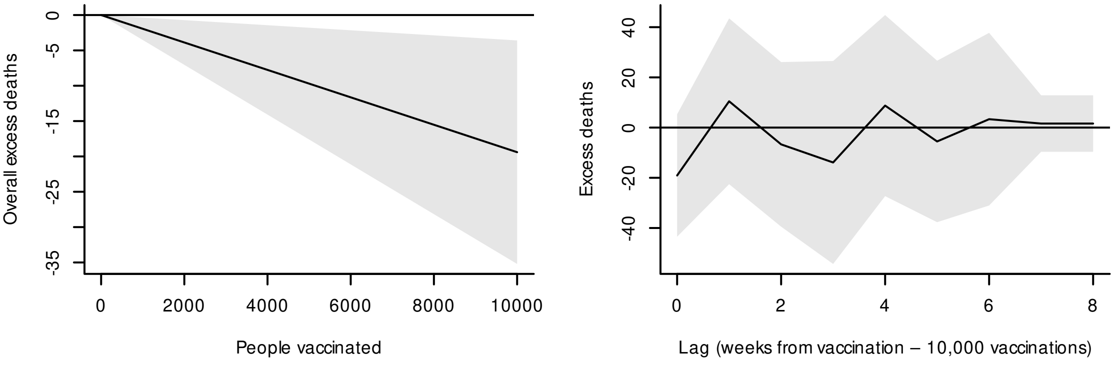


**Ages 70+ ages, long cross-basis (DLNM)**

Estimate: 27.65 (95% CI: -3.25 to 58.56) attributable excess deaths per 10,000 vaccinations
Also, 1.14 (95% CI: 0.79 to 1.48) attributable excess deaths per COVID-19 death

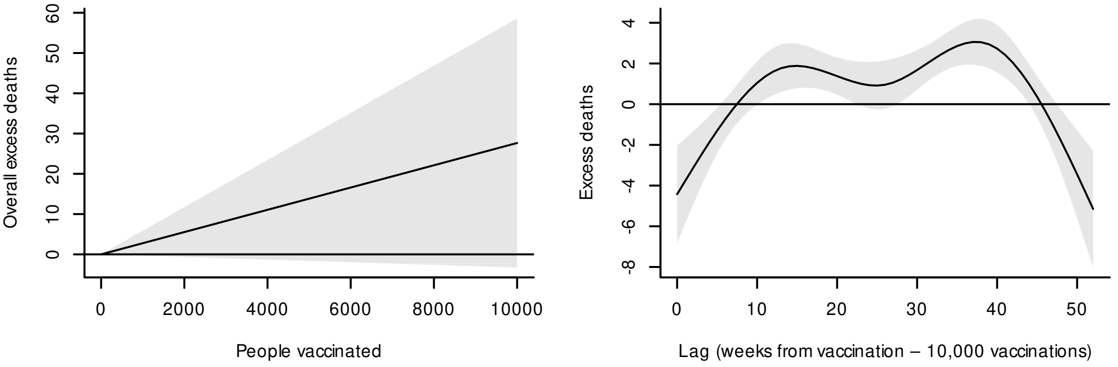


**Ages 70+ ages, long cross-basis (EuroMOMO)**

Estimate: 39.10 (95% CI: 3.24 to 74.96) attributable excess deaths per 10,000 vaccinations
Also, 1.03 (95% CI: 0.63 to 1.43) attributable excess deaths per COVID-19 death

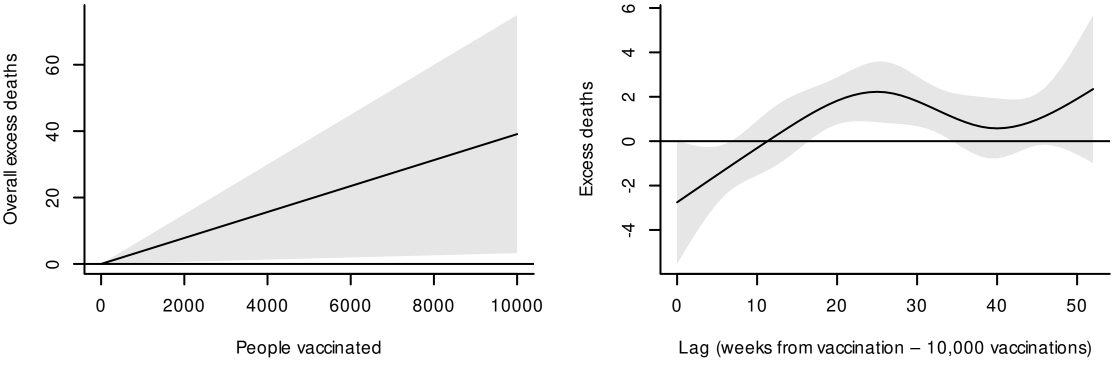
